# *STK11* mutations and deletions define a distinct subtype of cervical adenocarcinoma

**DOI:** 10.64898/2026.01.01.25343162

**Authors:** Emma Robinson, Elisabeth Murphy, Anaseidy Albanez, Sonam Tulsyan, Hong Lou, Yi Xie, Weiyin Zhou, Qian Yang, Tawnjerae Joe, Tongwu Zhang, Jia Liu, Wen Luo, Dongling Wu, Herbert Higson, Mathias Viard, Mary Carrington, Matt Oberley, Douglas R. Stewart, Hesler Morales, Lisa Mirabello, Enrique Alvirez, Roberto Orozco, Eduardo Gharzouzi, Michael Dean

## Abstract

Between 10 and 20% of cervical cancers are adenocarcinomas with poorer five-year survival and higher recurrence. To identify somatic alterations driving cervical cancer, we performed whole-exome sequencing of 308 subjects with invasive disease from Guatemala and Venezuela. Consistent with other studies, there is a higher rate of *TP53* mutations in adenocarcinomas versus squamous cell carcinomas (SCC), especially in HPV-negative tumors. We identified a higher rate of mutations and deletions (23%) in the *STK11* tumor suppressor gene in adenocarcinomas versus SCC. This result was confirmed in the AACR Project Genie and Caris cohorts. Whole-genome sequencing and SNP-array data identified significant numbers of focal deletions on chr19p that disrupt *STK11*, undetected by exome sequencing. In one tumor, HPV integration disrupts *STK11*. Chr19p is commonly deleted in cervical cancer, and we document a high rate of independent inversions, chromosomal translocations, and breakage-fusion-bridge events that provide the second hit to *STK11*. Significantly, *STK11* alterations are associated with a younger age of onset and poorer overall survival and survival on immunotherapy. Apart from *STK11*, *PIK3CA* mutations and *YAP1* amplification are prevalent cervical cancer drivers. *STK11* mutations and deletions co-occur significantly with YAP1 amplifications, suggesting an interaction between these pathways. In contrast, STK*11* alterations are mutually exclusive to *PIK3CA* mutation, suggesting redundancy. Cervical adenocarcinomas exhibit significantly lower *CD274* (PD-L1) expression and a poorer response to immune checkpoint inhibitors (ICIs). *STK11* mutations are associated with poor responses to ICI in other cancers, and elucidating the role of STK11 in cervical cancer may improve targeted and immunotherapies.

**Significance:** Characterized one of the largest cohorts of fresh-frozen, invasive cervical cancer cohorts and documented that *STK11* mutations and deletions are more prevalent in adenocarcinoma, and defined an early-onset, more aggressive subtype.

## Introduction

Cervical cancer is one of the most common cancers in women globally, accounting for over 600,000 new cases and 340,000 new deaths in the year 2020 [1]. Over 90% of cases and deaths occur in low- and middle-income countries [2]; however, few studies of cervical cancer have been conducted in these populations. Although Guatemala has one of the highest rates of cervical cancer in the world, there are few studies of invasive cervical cancer in the country [2]. Characterizing the disease in low and middle-income countries (LMICs) is therefore critical to reducing disease incidence and mortality.

Histologically, cervical cancer is defined by two major subtypes: adenocarcinoma (AD) and squamous cell carcinoma (SCC). Most cervical cancer cases are SCC (83%), and the two subtypes are epidemiologically and prognostically distinct [3]. While HPV16 is the most common driver of both subtypes, HPV18 is found more frequently in AD than SCC, and distinct sublineages of HPV16 confer elevated risk of AD [4, 5]. AD patients are frequently diagnosed at a younger age than SCC patients with more advanced disease and have poorer five-year survival and higher recurrence [6, 7]. Cytology is less effective at detecting AD than SCC, leading to diagnoses at more advanced stages [8].

Despite the heavy burden of disease, cervical cancer treatments is mainly confined to surgery, chemotherapy, and radiation. Whole-exome sequencing studies have identified the major somatic driver genes for cervical cancer including *PIK3CA*, *EP300*, *FBXW7*. *PTEN*, and *HLA-A* [9-11]. However, there are few approved therapies for cervical cancer targeted to specific subtypes or genetic variants, as there are for other cancers such as breast cancer. We have previously reported on two frequent and mutually exclusive cervical drivers: *PIK3CA* mutation, which is present in 32% cervical cancer cases, and *YAP1* amplification, which is present in 12% of tumors and associated with poorer survival [12]. Additionally, we and others have demonstrated that a PI3K inhibitor approved to treat breast cancer is effective in impeding the growth of *PIK3CA*-mutated cell lines [13] (manuscript submitted). However, more research is needed to further characterize the genetic factors driving cervical cancer to develop targets for more effective therapies.

To identify additional genetic drivers of cervical cancer, we obtained 306 invasive cervical cancer tumors from patients in Guatemala and Venezuela, with matching blood samples for 226 patients: the Latin American Cervical Cancer Cohort. We performed whole-exome sequencing to determine the alterations of previously identified cancer driver genes and investigated associations between histology, HPV type, and age and gene-alteration frequencies.

## METHODS

### Patient Recruitment

Primary, and treatment naïve tumors and matching blood samples were collected from women (21-86 years of age) evaluated for cervical cancer, before therapy, at the Instituto Nacional de Cancerologia (INCAN) in Guatemala and Hospital Central Universitario (Dr. Antonio Maria Pineda) in Venezuela from 2011 to 2015. The IRBs at the Guatemalan and Venezuelan hospitals approved the study, and all patients provided informed consent. Trained personnel completed questionnaires with reproductive and risk factor variables. Data on weight or BMI was not obtained. All samples were coded. All samples were collected prior to the women receiving nonsurgical treatment.

### Whole Exome Sequencing

Targeted sequencing was performed on randomly selected samples, based on DNA availability. of tumor and normal (blood) samples of CESC patients was conducted to identify somatic variants. DNA libraries were created from 200 ng of tumor or blood DNA using the Kapa HyperPlus Kit and xGen Dual Index UMI Adapters, according to the protocol provided by Kapa. Each multiplexed sequencing library was then sequenced on the Illumina NovaSeq 6000 using the paired-end 200-bp strategy. Read lengths of 2 x 150 bp were used, yielding an average coverage of 500X for tumor samples and 50X-100X for normal (blood) samples. Samples were quality controlled, alignment to the human hg38 build of the genome, and variant calling performed by Manta, Muse, Mutect2, Strelka, and Vardict. Variants that occurred in both tumor and normal DNA across the paired samples were excluded, and a panel of normal was used to filter tumor-only samples.

### Verification of Mutational Data

Mutational data for 315 genes known to be frequently mutated in cancer [14] were analyzed further and a subset of 24 of these genes was designated as “cervical cancer driver genes” based on previous research [9-11]. Each mutation found in the 315 cancer genes was further evaluated to determine if it was true or an artifact. For tumor only analysis, mutations were excluded if they met any of the following conditions: 1) having an overall allelic frequency of greater than 0.01% in gnomAD v 4.1 [15]; 2) being designated as “Benign” or “Likely Benign” in ClinVar [16]; 3) having a variant allele frequency (VAF) of less than 2% from WES mutation calling; and 4) were not found as somatic mutations in tumor-normal pairs but observed in multiple tumor-only samples. For tumor-normal pair analysis, mutations were excluded from the dataset if they were designated as “Benign” or “Likely Benign” in ClinVar (RRID:SCR_006169) [16].

### HPV Typing

Cervical tumor DNA was amplified with the gp5/6 HPV L1 gene primers and Sanger sequenced. Sequences were used in a BLAST (RRID:SCR_004870) search to identify the HPV type. In addition, selected samples were sequenced with an Ion Torrent overlapping primer set targeting 14 HPV types [17]. Not all samples contained sufficient DNA for HPV typing after whole-exome sequencing. To fill in missing HPV type data and validate HPV typing conducted using the above methods, we aligned exome sequence data to 65 HPV types. Because this method is less sensitive than other methods, only samples with confirmed integration sites, or greater than 100 viral reads were called positive, and ambiguous samples were labeled “unknown” rather than “negative.” Samples labeled as negative were negative in one or both PCR-based assays.

### Mutational Signature Analysis

Somatic SNVs from VCF files were processed using SigProfilerMatrixGenerator (Python, RRID:SCR_024202) to generate single-base substitution (SBS96) mutational count matrices based on the human reference genome (GRCh37). SNVs were categorized into 96 trinucleotide contexts following standard COSMIC (RRID:SCR_002260) definitions. De novo mutational signatures were extracted from the SBS96 matrix using SigProfilerExtractor (RRID:SCR_023121) with default NMF-based workflows. A range of potential signature numbers was evaluated, and the final solution was automatically selected based on signature stability, reproducibility, and sample reconstruction accuracy. Extracted signatures were then compared with COSMIC v3 reference mutational signatures using cosine similarity, and each de novo signature was assigned to its closest COSMIC match. Signature exposures were estimated per sample and normalized to the total mutation burden. Samples with insufficient SNV counts were excluded from de novo extraction to ensure robust signature assignment. Some samples originally classified as HPV-negative had strong APOBEC signatures and were designated as HPV “hit and run” samples [18].

### Structural Variant calling of Latin American tumors

Amplifications and deletions of specific gene regions were determined using the allelic ratio of variants from either the WES or SNP array data. The ASCAT (RRID:SCR_016868), GISTIC (RRID:SCR_000151), and MoChA programs were used, and the analyses incorporated results from all three methods. ASCAT derives copy-number profiles of tumor cells, accounting for normal cell admixture and tumor aneuploidy, thereby inferring tumor purity and ploidy [19]. GISTIC identifies regions of the genome that are significantly amplified or deleted, and was applied to tumor-normal pairs as well as tumor-only samples [20]. MoChA calls mosaic chromosomal alterations starting from phased VCF files [21]. Targeted analysis was performed for amplification of the *YAP1*, *CD274* (PD-L1), *EGFR*, and *ERBB2* (HER2) genes and deletions of *STK11* and *PTEN*.

### Focal deletions in the *STK11* gene in WGS

Focal deletions in cervical cell lines (6 in total) and 14 TCGA (RRID:SCR_016637) CESC samples were identified by manual review in IGV of WGS. Junction reads were used to determine the breakpoints of the deletions. The copy number of selected chr19 genes was used to identify TCGA tumors with 19p copy number loss. Manual review of WGS data was used to determine the breakpoint of deletion events. Putative breakage-fusion-bridge (BFB) events were identified by the characteristic stair-step pattern of read depth and the presence of inverted reads spaced 500-2000 bp apart due to digestion and deletion of the remainder of the chromosome to the telomere [12].

### Validation in publicly available datasets

The TCGA, AACR Project GENIE (AACR), and MSKCC data were accessed via cBioPortal (<u>cBioPortal for Cancer Genomics</u>) [22]. Data on Japanese cervical cancer are from Hirose 2020 [23].

#### Caris Data

This retrospective study was conducted under Caris Life Sciences’ Research Data Banking protocol, which was reviewed and granted IRB exemption by the WCG IRB. The study adhered to the ethical guidelines of the Declaration of Helsinki, the Belmont Report, and the U.S. Common Rule 45 CFR 46.104(d)(4).

### Caris-Next-Generation DNA Sequencing

NGS was performed on genomic DNA isolated from formalin-fixed paraffin-embedded (FFPE) tumor samples using the NextSeq or NovaSeq 6000 platforms (Illumina, Inc., San Diego, CA). For NextSeq-sequence tumors, a custom-designed SureSelect XT assay was used to enrich for 592 whole-gene targets (Agilent Technologies, Santa Clara, CA). For NovaSeq-sequenced tumors, more than 700 clinically relevant genes were sequenced at high coverage and read depth, along with an additional panel designed to enrich for>20,000 genes at lower depth. All variants were detected with >99% confidence based on allele frequency and amplicon coverage, with an average sequencing depth of >500 and an analytic sensitivity of 5%. Before molecular testing, tumor enrichment was achieved by manual microdissection of targeted tissue. Genetic variants identified were interpreted by board-certified molecular geneticists and categorized as ‘pathogenic,’ ‘likely pathogenic,’ ‘variant of unknown significance,’ ‘likely benign,’ or ‘benign,’ according to the American College of Medical Genetics and Genomics (ACMG) standards. When assessing mutation frequencies of individual genes, ‘pathogenic’ and ‘likely pathogenic’ were counted as mutations. The copy number alteration (CNA) of each exon is determined by calculating the average depth of the sample and the sequencing depth of each exon and comparing the computed result to a pre-calibrated value.

### Caris-Whole Transcriptome Sequencing

Gene fusion detection was performed on mRNA isolated from a formalin-fixed paraffin-embedded tumor sample using the Illumina NovaSeq platform (Illumina, Inc., San Diego, CA) and Agilent SureSelect Human All Exon V7 bait panel (Agilent Technologies, Santa Clara, CA). FFPE specimens underwent pathology review to determine tumor content and tumor size; a minimum of 10% tumor content in the microdissection area was required to enable enrichment and extraction of tumor-specific RNA. The Qiagen RNA FFPE tissue extraction kit was used for extraction, and the RNA quality and quantity were determined using the Agilent TapeStation (RRID:SCR_019547). Biotinylated RNA baits were hybridized to the synthesized and purified cDNA targets, and the bait-target complexes were amplified in a post-capture PCR reaction. The resultant libraries were quantified, normalized, and the pooled libraries were denatured, diluted, and sequenced; the reference genome used was GRCh37/hg19 and analytical validation of this test demonstrated ≥97% Positive Percent Agreement (PPA), ≥99% Negative Percent Agreement (NPA) and ≥99% Overall Percent Agreement (OPA) with a validated comparator method.

### Caris-RNA expression

FFPE specimens underwent pathology review to measure tumor content and tumor size; a minimum of 10% tumor content in the microdissection area was required to enable enrichment and extraction of tumor-specific RNA. Qiagen RNA FFPE tissue extraction kit was used for extraction, and the RNA quality and quantity were determined using the Agilent TapeStation. Biotinylated RNA baits were hybridized to the synthesized and purified cDNA targets, and the bait-target complexes were amplified in a post-capture PCR reaction. The Illumina NovaSeq 6500 was used to sequence the whole transcriptome from patients to an average of 60M reads. Raw data were demultiplexed using the Illumina DragonBio IT Accelerator, trimmed, counted, PCR duplicates removed, and aligned to the human reference genome hg19 using the STAR aligner. For transcription counting, transcripts per million values were generated using the Salmon expression pipeline.

### Caris-HPV16/18 typing by WES

HPV16/18 was detected using the Caris pipeline, which includes 39 unique baits to detect HPV16 and 50 unique baits to detect HPV18 out of a total of 2360 pathogen baits. The threshold for positivity is ≥ 100 reads for either HPV16 or HPV18.

### Caris-Tumor Mutational Burden (TMB)

TMB was measured by counting all non-synonymous missense, nonsense, in-frame insertion/deletion, and frameshift mutations found per tumor that had not been previously described as germline alterations in dbSNP151, Genome Aggregation Database (gnomAD)databases, or benign variants identified by Caris geneticists. A cutoff point of >=10 mutations per MB was used based on the KEYNOTE-158 pembrolizumab trial [24], which showed that patients with a TMB≥ 10 mt/MB across several tumor types had higher response rates than those with a TMB < 10 mt/MB. Caris Life Sciences is a participant in the Friends of Cancer Research TMB Harmonization Project [25].

### Caris-Microsatellite instability (MSI)-WES

Microsatellite instability was assessed using NGS by directly analyzing 2,810 known homopolymers in pentapolymer target microsatellite regions sequenced in the WES gene panel, and by comparing the resulting sequence reads with the reference genome hg38 in the University of California, Santa Cruz (UCSC) Genome Browser database. The number of microsatellite loci that were altered by somatic insertion or deletion was counted for each sample. Only insertions or deletions that result in an increase or decrease in the number of tandem repeats were considered. Genomic variants at microsatellite loci were detected using the same depth and frequency criteria as for mutation detection. The threshold for MSI high (MSI-H) by NGS was determined to be 116 or more loci with insertions or deletions; equivocal to be 113-115, while stable to be 112 or less.

### Caris-Overall Survival

Overall survival (OS) was defined as the time from the index date (sample collection) to death or last clinical contact. For patients with a recorded date of death, OS was calculated as the number of days from sample collection to death. For patients without a recorded date of death, OS was calculated as the number of days from sample collection to the last clinical contact, defined as the maximum date observed across all claims sources and the collection date. Patients without a recorded death date were classified as having an inferred death if their last clinical contact occurred more than 100 days before the global last contact date in the dataset. If there was no recorded or inferred death, then patients were censored at the date of last clinical contact.

### Caris-Time on Treatment (ToT)

Time on treatment (ToT) for ICI drugs was defined as the time elapsed between the recorded drug start date and the drug end date. Patients without a record of the drug of interest were excluded from ToT analyses. Censoring status for ToT was assigned based on treatment discontinuation and death. Patients were considered uncensored if a drug end date was observed, if a recorded or inferred death occurred, or if ongoing treatment was not confirmed within 100 days of the most recent data refresh. Patients were considered censored if no recorded or inferred death was present and treatment continuity was confirmed within 100 days of the most recent data refresh, indicating ongoing therapy at the time of data cutoff.

Clinical outcomes statistical analysis: Overall survival and time on treatment were estimated using the Kaplan-Meier method and compared across defined cohorts of the analysis population. Survival and treatment curves are visualized with 95% confidence intervals, and risk tables are generated to display patient numbers and events over time. Differences in survival between defined cohorts were examined using Cox proportional hazards regression to estimate hazard ratios (HRs).

### Statistical Analyses

Chi-squared and Fisher’s exact tests were used to investigate differences in mutational frequency by histology and HPV type for each gene. Associations between driver genes were also examined using chi-squared tests and Fisher’s exact tests. Wilcoxon tests were conducted to investigate differences in mutational frequencies by age. Results were validated using the Caris, TCGA, and AACR datasets. RStudio (R version 4.4.0) was used to conduct selected analyses.

### Data Availability

Data will be deposited in dbGAP under accession ID xxx.

## Results

### Demographics

Histological data were available for 68% of the 306 Latin American samples. Over half of the samples (52%) were squamous cell carcinoma (SCC), while 16% were adenocarcinoma (AD). The median age of the cohort was 51.5 years (IQR 44.0-61.0) (**Table 1, Supplemental Figure 1**). The median age differed little between individuals with SCC (51.2) and individuals with AD (51.8). Two rare soft tissue sarcomas of the cervix were also ascertained- one sample each with leiomyosarcoma and rhabdomyosarcoma. The most common HPV type among the samples was HPV16 (29%), followed by HPV18 (18%) and HPV45 (12%). HPV16 is more frequent and HPV18 and 45 are less frequent in SCC compared to AD (HPV16 33% in SCC, 17% in AD; HPV18/45 24% and 20% in SCC, 31% and 23% in AD (**Table 1**).

**Table 1.**
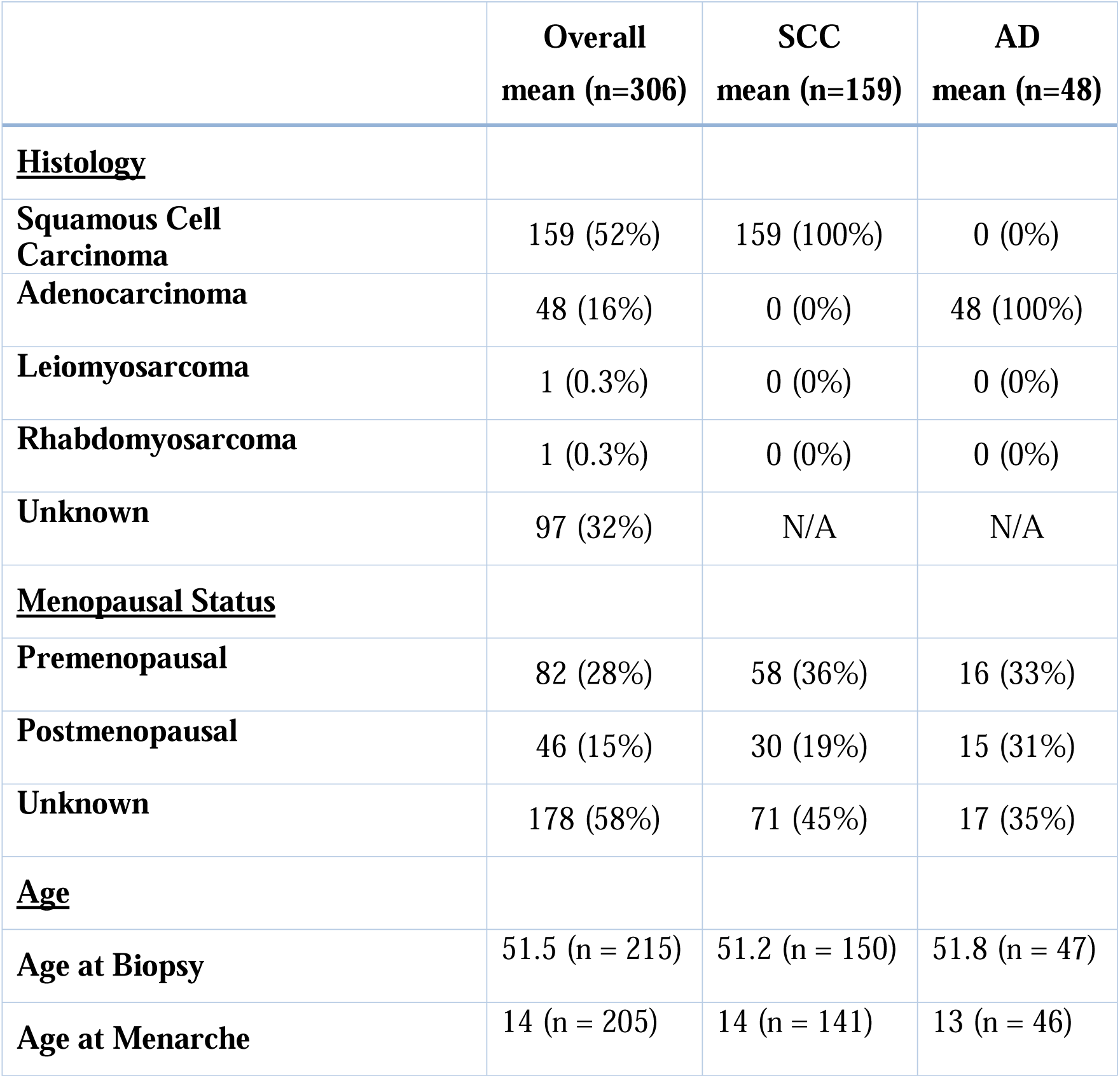
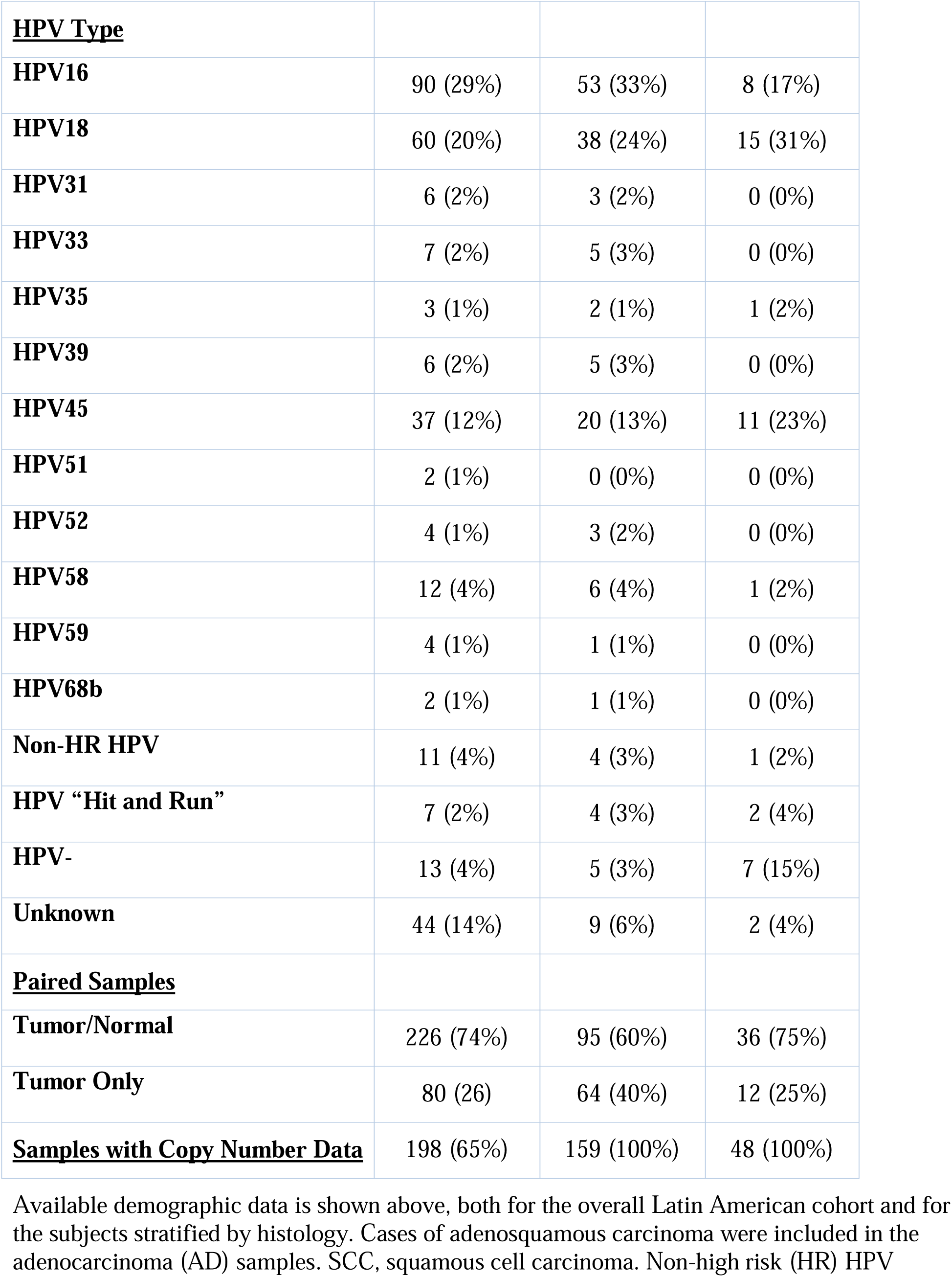
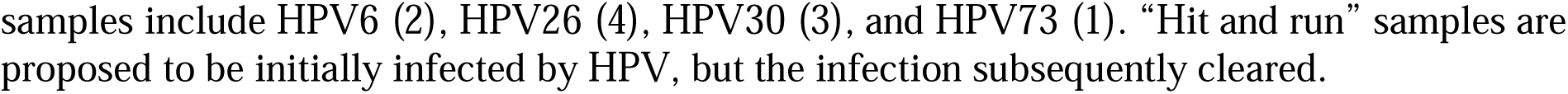
Demographics of the Latin American Cohort.

Available demographic data is shown above, both for the overall Latin American cohort and for the subjects stratified by histology. Cases of adenosquamous carcinoma were included in the adenocarcinoma (AD) samples. SCC, squamous cell carcinoma. Non-high risk (HR) HPV samples include HPV6 (2), HPV26 (4), HPV30 (3), and HPV73 (1). “Hit and run” samples are proposed to be initially infected by HPV, but the infection subsequently cleared.

### Cancer Driver Gene Mutational Analysis

To identify the most frequently mutated driver genes in Latin American cervical cancer, we performed whole-exome sequencing on 306 samples (**Figure 1; Supplemental Figure 2**). We assessed the frequency of mutations and copy-number variations in cancer driver genes (14). As previously described, the most frequently mutated gene was *PIK3CA* (35%) (10). Other frequently mutated driver genes included *FAT1* (18%), *FBXW7* (12%), and *STK11* (10%)(**Supplemental Table 1**). A whole genome copy number analysis identified amplification and deletion regions of the genome, including gain of chr3q (*PIK3CA*/*TP63*), chr7p11 (*EGFR*), chr9p24 (*CD274*/PD-L1), chr11q *YAP1*/*BIRC2*/*BIRC3*), chr17q12 (*ERBB2*), and loss of 19p (*STK11*) (**Supplemental Figure 3**). Analysis of the gain or loss of known drivers showed that 14% of tumors had *STK11* deletion, whereas 5% of individuals had YAP1 amplification (**Supplementary Figure 4**).

**Figure 1.**
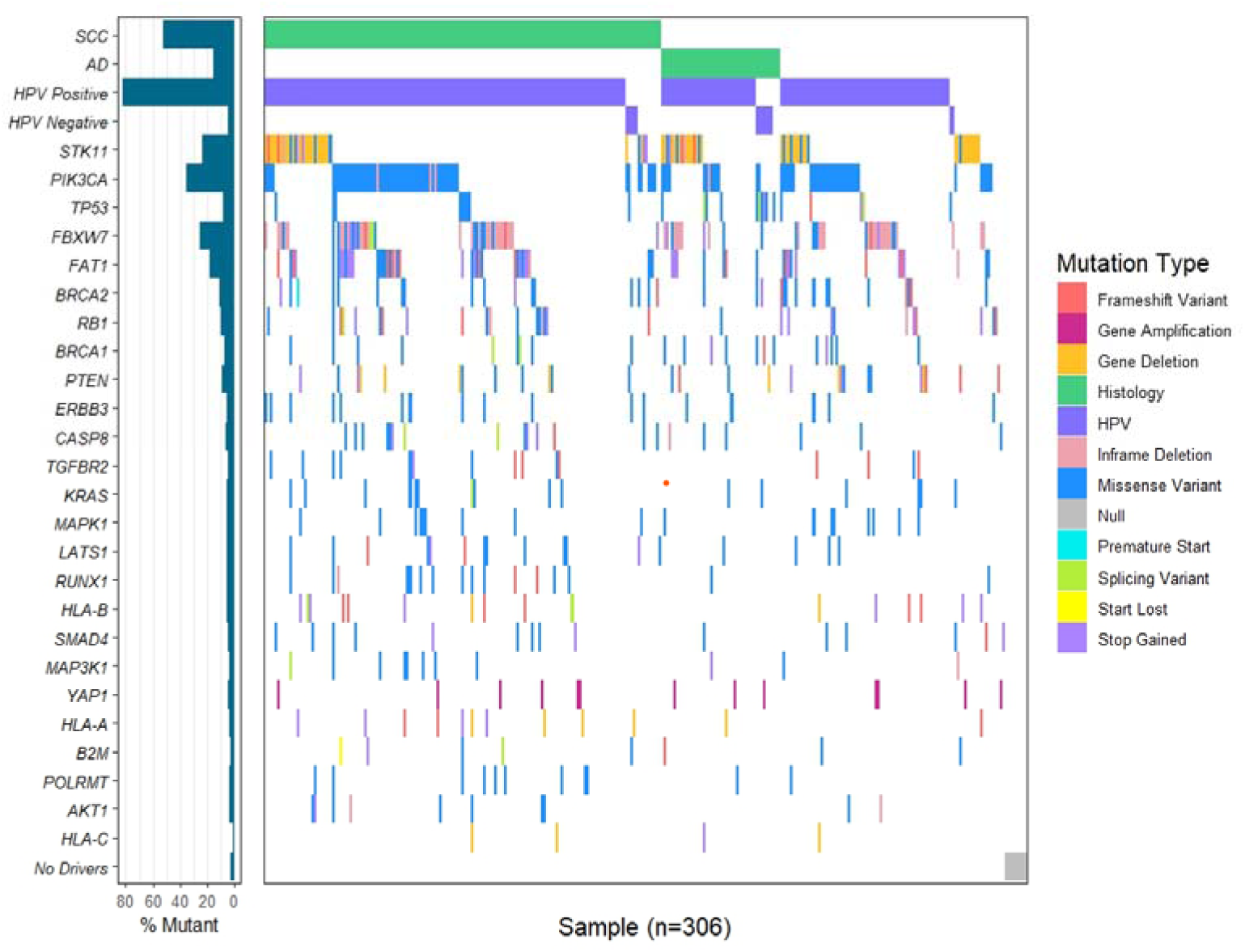
Waterfall plot of histology, HPV status, and somatic driver mutations in Latin American cervical cancer. Somatic mutations in 24 cervical cancer driver genes (**Supplementary Table 1**) are shown for squamous cell carcinoma (SCC), adenocarcinoma and adenosquamous cell carcinoma (AD)(purple), and unknown histology (blank). The *YAP1* line represents gene amplification; all others are mutations only. Individual SCC and AD plots are in **Supplemental Figure 2**.

### Comparison of driver gene variants in cervical AD and SCC

To identify genetic differences in the two most common histological subtypes of cervical cancer, the frequencies of driver gene mutations in adenocarcinoma and SCC were compared (**Figure 2A**). Two genes (*TP53* and *STK11*) were mutated significantly more frequently in adenocarcinoma than in SCC. Significantly more individuals with adenocarcinoma (21%) had a *TP53* mutation than individuals with SCC (6%)((X^2^=4.4; p = 0.0037). In adenocarcinomas, 23% of tumors harbored a *STK11* mutation, compared with only 7% of individuals with SCC (p = 0.0039). The association between *TP53* and adenocarcinoma has been described previously [26]but the association between *STK11* mutation and cervical adenocarcinoma has not been investigated and warrants further investigation. As mentioned above, *STK11* was also frequently deleted in our dataset. The frequency of *STK11* deletions by histology trended in the same direction as mutations: 20% of individuals with adenocarcinoma had a *STK11* deletion, versus 12% in SCC (p = 0.20). In total, 23% of individuals with cervical cancer in our dataset had an alteration in *STK11*: 35% of individuals with adenocarcinoma had an alteration in *STK11* compared with 19% of individuals with SCC (p = 0.026). These findings suggest that alterations to *STK11* are one of the top genetic drivers of cervical cancer, along with *PIK3CA* mutations and *YAP1* amplification. In fact, *STK11* alterations are the most prevalent driver mutations in cervical adenocarcinoma in this dataset.

**Figure 2.**
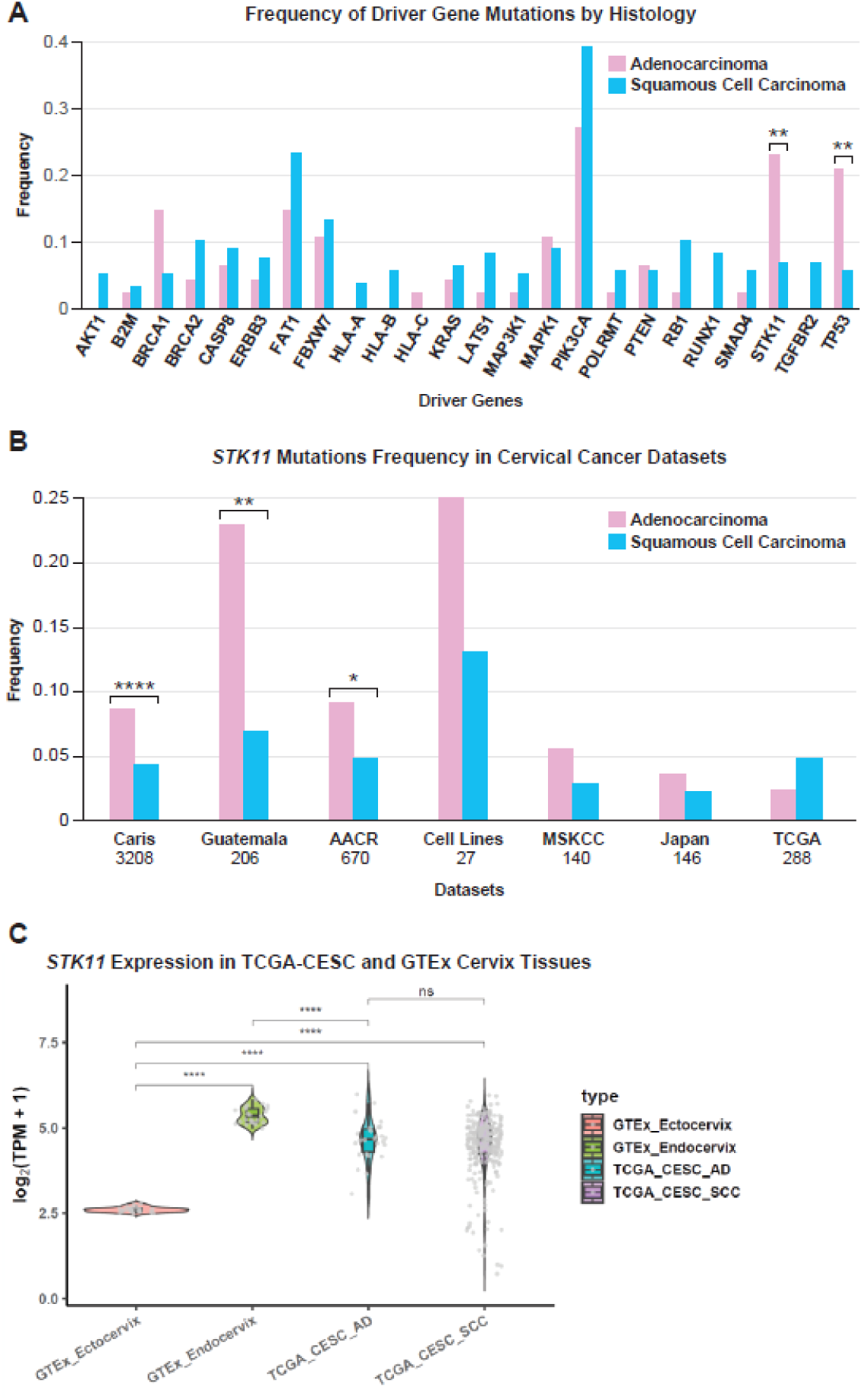
*STK11* is a significant driver of cervical adenocarcinoma. **A.** The frequency of somatic driver mutations in Latin American cervical cancer is shown, comparing adenocarcinoma and squamous cell carcinoma. **B,** The frequency of *STK11* mutations in different studies is shown with the sample sizes below the study name. **C,** The expression of the *STK11* gene is compared in normal cervical ectocervix and endocervix from GTEx, and compared to adenocarcinoma (AD) and squamous cell carcinoma (SCC) in TCGA. *=P<0.01, **=P<0.001, ***=P<0.0001, ****=P<0.00001.

The higher prevalence of *STK11* mutations and cervical adenocarcinoma was validated in other datasets. In the AACR dataset, 8.3% of 325 individuals with adenocarcinoma and 4.3% of 345 individuals with SCC had an *STK11* mutation (p = 0.035). In the Caris CodeAI dataset, 8% of 828 individuals with adenocarcinoma and 4% of 2380 individuals with SCC had an *STK11* mutation (p = 2.5e-06). This trend held but was not significant in a Japanese dataset [23], the MSKCC dataset [27], and our cervical cell line data. The only dataset in which *STK11* mutations occurred more frequently in SCC was TCGA, but this difference was not significant (**Figure 2B**). In the Caris CodeAI dataset, *STK11* deletions occurred significantly more frequently in adenocarcinoma (9.0%) than SCC (4.7%)(p = 7e-06). This same trend was observed but was not significant in a Japanese dataset (22), the MSKCC dataset, and a panel of 28 cervical cancer cell lines. *STK11* deletions occurred slightly more frequently in SCC in the TCGA and AACR datasets, but this difference was not significant (**Figure 2B, Supplemental Figure 5**). Interestingly, the expression of *STK11* is very low in normal cervical ectocervix, the precursor to adenocarcinomas (**Figure 2C**).

To study the impact of large deletions of chr19p, including *STK11*, we analyzed whole-genome sequence (WGS) data from 27 cervical cancer cell lines. We identified six cell lines with deletions ranging from 8.4 to 100.4 KB, deleting all or part of the *STK11* gene (**Figure 3A, Supplemental Table 2).** We searched for similar deletions in 270 TCGA cervical cancers with WGS. Whereas the TCGA PanCancer analysis identified seven cases of homozygous deletion using GISTIC, our study identified 13 additional tumors with either homozygous or focal deletions in *STK11* (**Figure 3B**). The focal deletions ranged from 41 bp to 1.1 MB (**Supplemental Table 2**). GISTIC values for genes on chr19p were investigated to determine the mechanism of deletion. Examination of copy number values for genes on chromosome 19p showed that low copy number for *STK11* was observed in 12 samples, with a region of focal high copy number values centromeric to *STK11*, consistent with breakage-fusion-bridge events (**Supplemental Figure 6, Supplemental Table 3**). Long read WGS allowed the resolution of a BFB event at 10.3 MB in the SNU-902 cervical cancer cell lines (**Supplemental Figure 6).** The site of the BFB events ranged from just centromeric to *STK11* (1.8 MB) to 43.5 MB, with a cluster of six BFBs from 7.5-8.5 MB. An additional 10 samples were identified, including inversions, rearrangements, and translocations of chr19p to other chromosomes (**Supplemental Figure 6**). The TCGA-VS-A8Q9 sample shows integration of HPV51 at chr19:12.79 MB, with a 10-fold amplification of *JUNB*, which may be due to ecDNA (**Supplemental Figure 7**). *STK11* deletions were identified in whole-genome SNP array data from Latin American cervical cancer samples using the MoChA program (20), and 17 tumors had focal loss of all or part of the *STK11* gene (**Figure 3, Supplemental Figure 4B**). Lastly, we compiled all identified mutations in the *STK11* gene in cervical cancers, and 82% are either loss-of-function or recurrent somatic. Interestingly, the S216F mutation is common across studies and is a TCC-to-TTC mutation consistent with APOBEC mutagenesis (**Figure 3D, Supplemental Table 4).**

**Figure 3.**
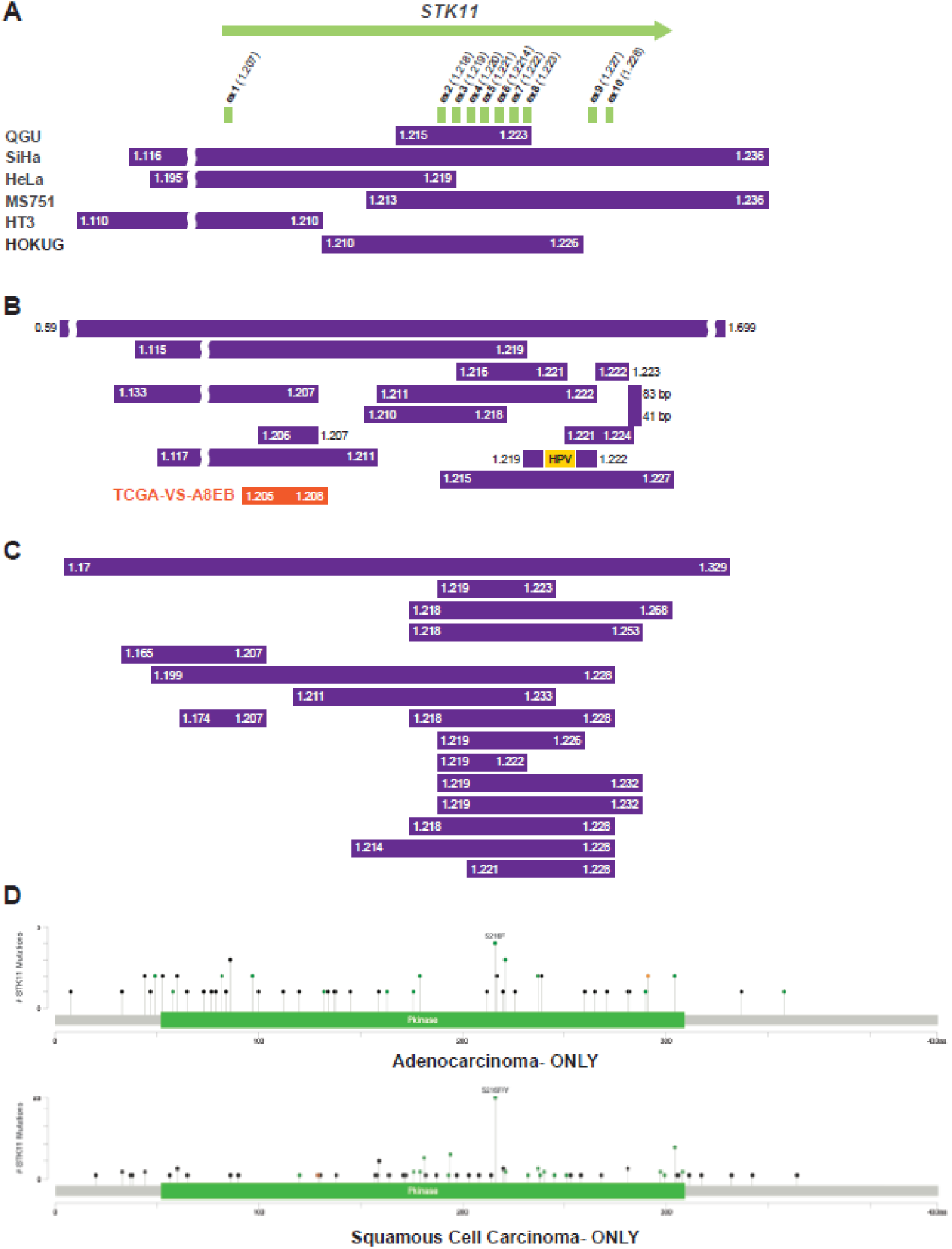
Focal deletions and mutations in cervical tumors. **A,** the position of focal deletions on chr19, including all or part of the *STK11* gene in 19p13.3, in six cervical cancer cell lines. All cell line deletions are homozygous with loss of the non-deleted allele (**Supplemental Figure 8**). The purple bars indicate the deleted region with the chr19 location in MB given at the start and end. **B,** focal deletions that were previously undetected in the WES analysis of the *STK11* gene are shown in individual TCGA cervical tumors. The yellow bar represents the location of HPV integration**. C,** focal deletions in the *STK11* gene in Latin American cervical cancer detected by MoChA analysis of SNP array data. **D,** plots of the cervical cancer *STK11* mutations in Caris samples separated by adenocarcinoma and squamous cell carcinoma. Ex=exon, del=deletion, HOMDEL=homozygous deletion. Green=missense, black=truncating, orange=splice site mutations.

To investigate the role of *STK11* mutations and deletions on the age at diagnosis, we examined AD and SCC tumors across multiple cohorts. In the Guatemala dataset, *STK11*-mutated individuals with AD had an average age of 42, significantly lower than that of *STK11-*WT individuals with AD (median age = *54;* p = 0.0083) (**Figure 4A).** Interestingly, there was no difference in the average ages of *STK11*-deleted individuals with AD (54) compared to *STK11-*WT individuals (54). In the Caris dataset, *STK11* alteration was associated with significantly younger age at collection in both SCC and AD (**Figure 4B)** and with lower *STK11* gene expression **(Figure 5).** This finding was further validated in the AACR dataset, where *STK11*-mutated individuals with AD had an average age of 44, and *STK11-*WT AD individuals had an average age of 49 (p = 0.029) (**Figure 4C**). Although *STK11* deletions trended in the same direction as mutations in the AACR Project Genie (AACR) dataset, *STK11*-deleted individuals with AD had an average age of 45, while *STK11*-WT individuals with AD had an average age of 49; this difference was not significant (p = 0.35) (**Supplemental Table 5**). In individuals with SCC in AACR, neither *STK11* mutations nor deletions were associated with age at diagnosis. These findings indicate that *STK11* mutation is associated with an earlier age of diagnosis, especially in individuals with AD.

**Figure 4.**
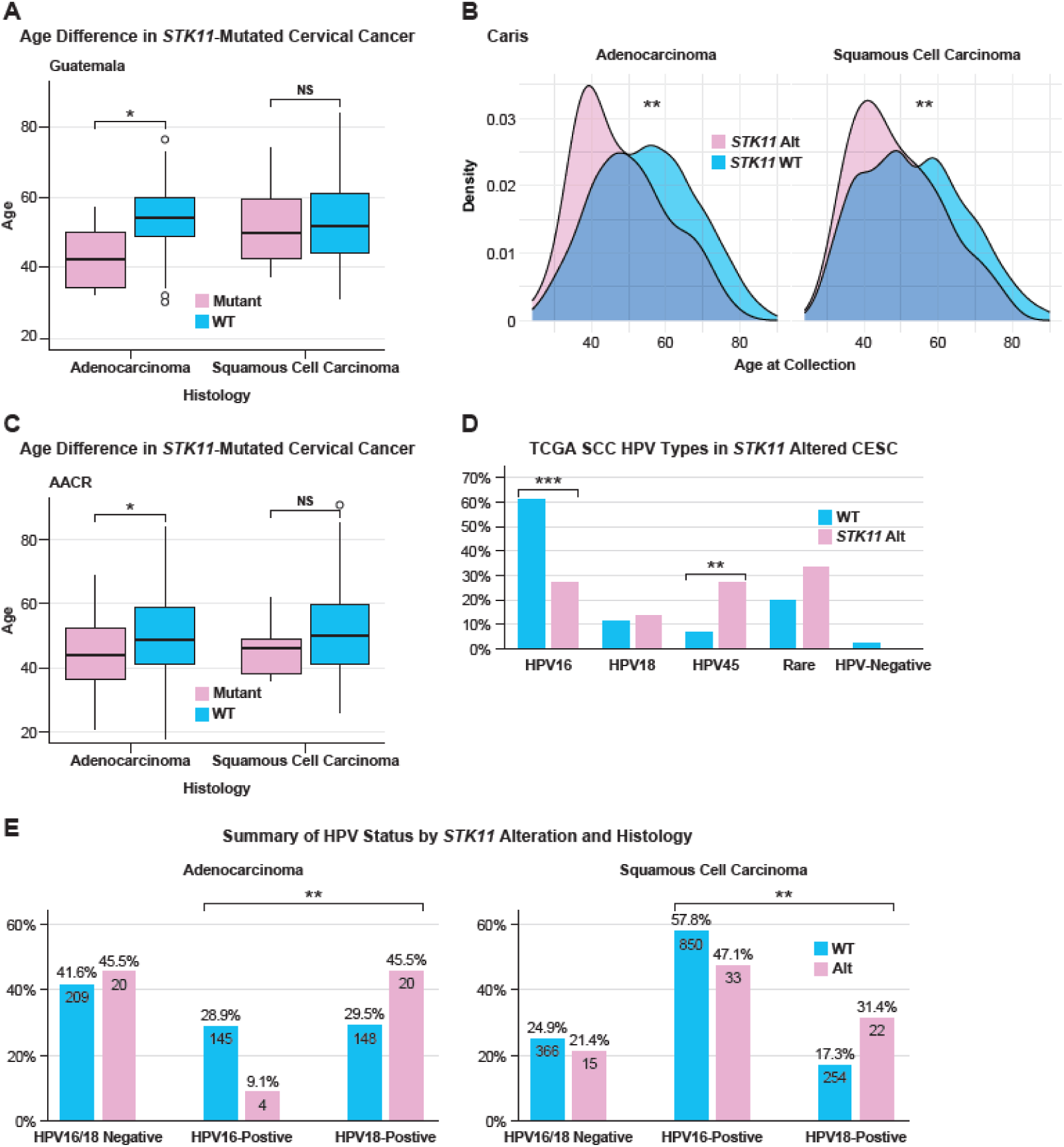
Difference in age of diagnosis, HPV type, and driver mutations in *STK11*-altered cervical cancer. **A**, The difference in age at collection by histological type is shown for Latin American cervical cancer. **B**, Caris samples displayed as curves, a box plot is in **Supplemental Figure 9E. C**, AACR Project Genie cervical cancer samples. **D**, The frequency of HPV types in adenocarcinoma (adeno) and SCC by *STK11* alteration status. **E**, the frequency of HPV16 and HPV18 by histological type and *STK11* status in the Caris cohort. WT, wild-type; Alt, altered (mutations and deletions)—significance values as in Figure 2.

**Figure 5.**
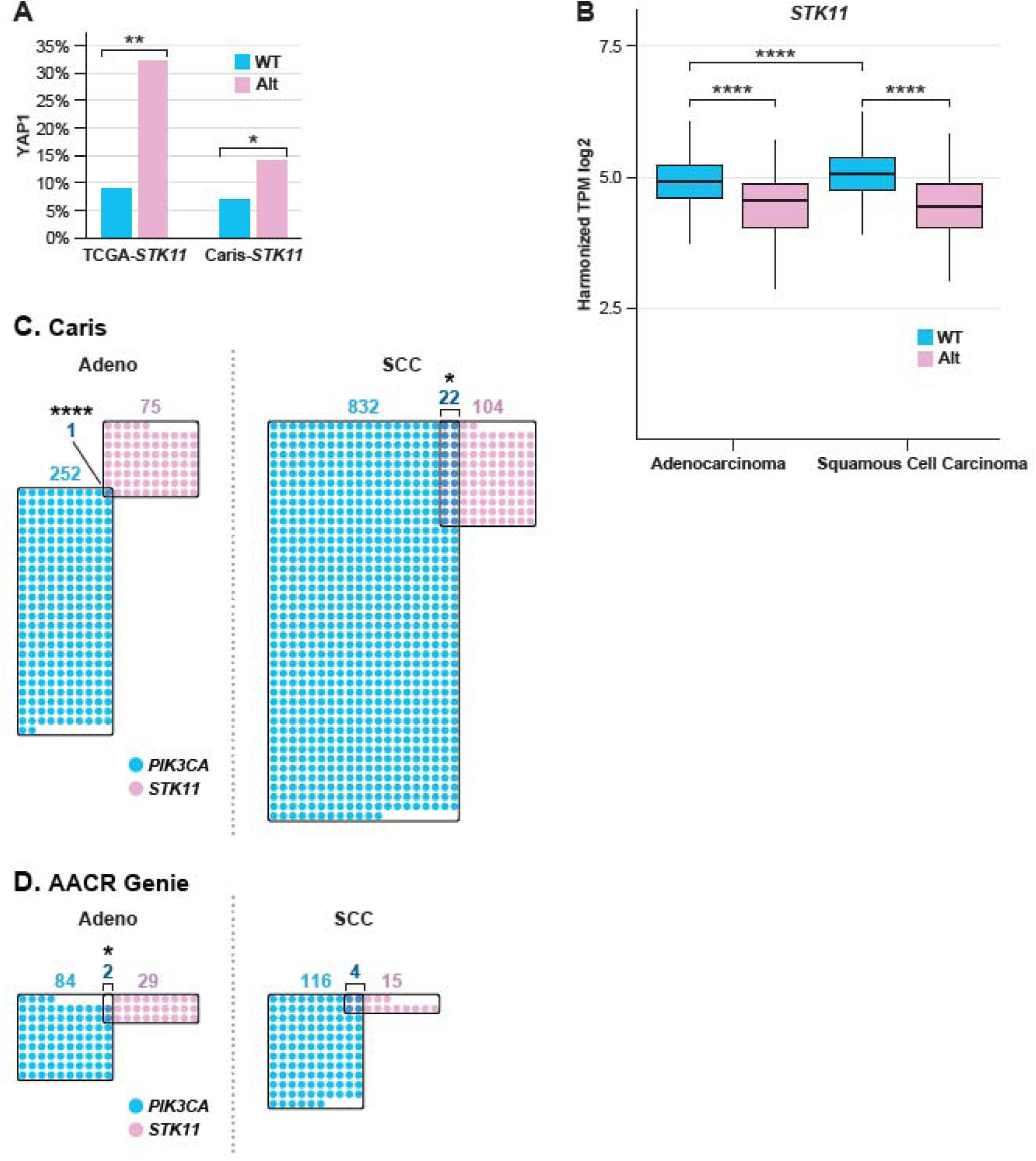
*STK11* alteration, *PIK3CA* mutation, and survival. **A,** The association of *YAP1*-amplification with *STK11* alteration in TCGA and Caris SCC. **B**, Normalized *STK11* expression is shown for Caris samples by histology and mutation status. **C,** The relationship between *PIK3CA* mutation and *STK11* alteration is TCGA samples shown for Caris **C,** and AACR. **D.** AMP-amplified, WT-wild-type, statistical significance as in Figure 2.

To determine the association between HPV type and *STK11* inactivation, *STK11* deletions and mutations were combined into a single *STK11*-alteration category in TCGA and Guatemala. In the TCGA dataset, HPV16+ SCC tumors were less likely to harbor an *STK11* alteration than those WT for *STK11* (p = 2.0e-04) (**Figure 4D**). Correspondingly, individuals who were HPV45+ were more likely to have an *STK11* alteration than those with STK11 WT tumors (p = 0.0028). In the Caris dataset, *STK11* alterations were less frequent in HPV16-positive samples and more frequent in *STK11*-WT samples across both adenocarcinoma and SCC (**Figure 4E**). Interestingly, no *STK11* mutations or deletions were observed in HPV-negative individuals (Figure 1; **Supplemental Figure 9A**). In Guatemala, no significant differences in HPV type frequency were observed; however, HPV45+ individuals were more likely to have *STK11* alterations than *STK11*-WT individuals (**Supplementary Figures 9B and 9C**).

### Association of *STK11* alteration with other drivers

We have previously established that two of the most frequent genetic alterations in cervical cancer (*PIK3CA* mutation and *YAP1* amplification) are mutually exclusive (12)(manuscript submitted). Considering this, we evaluated the frequency of co-occurrence between PIK3CA mutation or YAP1 amplification and *STK11* alteration. In all TCGA cervical tumors, 9% of *STK11*-WT tumors had *YAP1*-amplification, while 32% of *STK11*-altered tumors had *YAP1*-amplification (P=0.004)(**Figure 5A**). The same association is seen in Caris SCC, with 14% of STK-altered samples having *YAP1* amplification versus 7% in *STK11* wild-type samples (P=0.015)(**Figure 5A)**. In the AACR Project Genie and Caris datasets, *STK11* alterations are almost completely mutually exclusive with *PIK3CA* mutation (P<0.00001, in Caris) (**Figure 5C, D**). In SCC, *PIK3CA* mutation is also negatively correlated with *STK11* alteration, but to a lesser extent, and is significant only in Caris, Guatemala, and TCGA (**Supplemental Figure 10A**). In the Caris dataset, tumor mutation burden is significantly higher in SCC than in AD, as previously reported; however, *STK11* alteration is not associated with a higher mutation burden, and microsatellite instability (MSI) tumors are rare in both AD and SCC (**Supplemental Figures 10B and 10C**).

The impact of *STK11* alterations on survival was investigated in the Caris dataset, with adenocarcinomas without *STK11* alterations as the reference group. Individuals with *STK11*-mutated or deleted adenocarcinomas had poorer survival (HR=1.54, 95% CI 1.11-2.15) (p =0.011) (**Figure 6A**). In this dataset, patients with SCC have worse overall survival, with *STK11*-altered samples being the worst but not significantly worse than SCC *STK11* WT tumors (**Figure 6A**). We identified 1065 Caris cervical cancer subjects who received immune checkpoint inhibitor therapy, almost exclusively Pembrolizumab. *STK11*-altered adenocarcinoma subjects with *STK11* alteration had a poorer survival (HR 2.35, 95% CI 1.3-4.2, P=0.003)(**Figure 6B**). In the combined Caris cohort, dominated by SCC, neither *STK11 alteration nor PIK3CA mutation had a significant impact on survival; however, the uncommon STK11-altered/PIK3CA-mutated samples had poorer survival (HR 1.9; 95% CI 1.3-2.8;* P=0.002). We examined the effect of age at diagnosis and genetic ancestry on survival and found that patients diagnosed under 50 had a significantly lower hazard of death (HR=0.89). Individuals of African and East/South Asian ancestry had poorer survival. Therefore, we performed multivariate analysis, but the impact on the hazard ratio was minimal (**Supplemental Figure 11**. In conclusion, *STK11* alterations are associated with poorer survival overall in cervical cancer and in patients treated with ICI.

**Figure 6.**
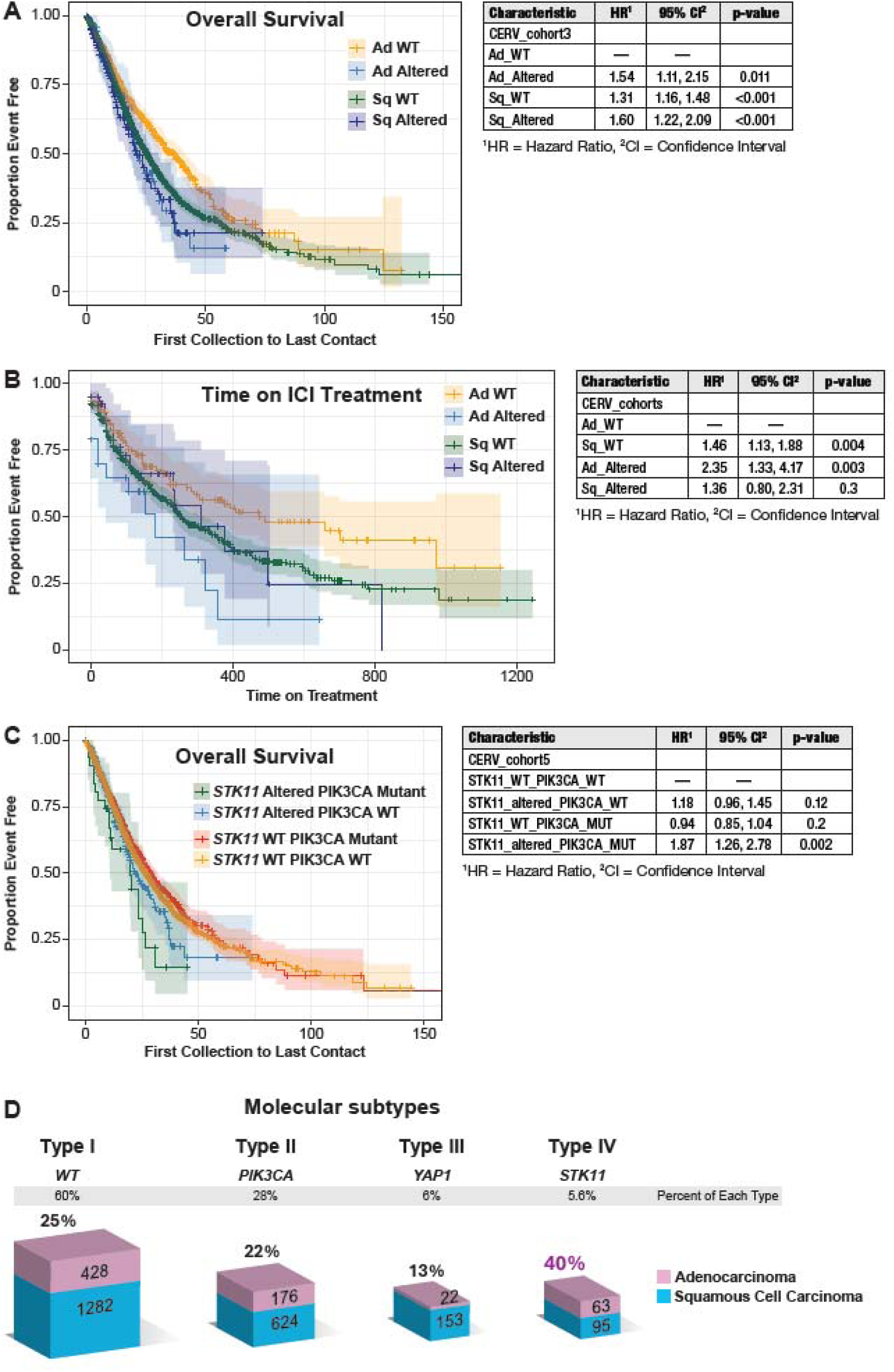
Survival in the Caris database of patients with *STK11*-altered tumors. **A.** Overall survival in months is shown for subjects from the first collection to the last contact for subjects with *STK11*-altered and wild-type (WT) tumors by histology (n=3790). **B.** Survival (days) of subjects treated with immune checkpoint inhibitors by *STK11* status and histology (N=1061). **C.** Combined survival (months) analysis of *STK11* alteration and *PIK3CA* mutation (N-3790). **D.** Bar graph of the four identified molecular subtypes and the percentage of AD. Sq, Squamous cell carcinoma, Ad, adenocarcinoma.

### Immune Escape Mutations

Mutations in genes involved in immune escape occurred relatively frequently in this sample: 16% of individuals had at least one mutation in *HLA-A*, *HLA-B*, *HLA-C*, *B2M*, or *CASP8*; a deletion in *HLA-A*, *HLA-B*, or *HLA-C*; or an amplification of *CD274/PD-L1*. The most frequently mutated immune gene was *CASP8*, with 7% of individuals carrying a *CASP8* mutation. In contrast, the least commonly mutated gene was *HLA-C*, with only 1 of 306 individuals carrying an *HLA-C* mutation (**Supplementary Figure 12)**. The frequency of immune escape mutations did not differ significantly between individuals with AD and those with SCC; however, *CD274* expression is significantly lower in TCGA AD compared to SCC (p = 0.0001) (**Supplemental Figure 13**).

### Rare Cervical Cancer Samples

Nearly all cervical cancer is driven by HPV. However, recent research has shown that 0.3% of cervical tumors are HPV-independent or negative, and these are predominantly AD with *TP53* mutations [28]. We initially identified 20 Latin American samples as HPV negative. Nine samples were AD (out of 48 total), nine were SCC out of (159 total), one was rhabdomyosarcoma, and one had unknown histology. Analysis of the mutational signatures of the tumors for which we had matching normal DNA allowed us to classify a subset of HPV-negative tumors as “HPV hit-and-run”. Although we were unable to detect HPV in these samples, based on the strong APOBEC signature of these tumors, it is likely that these individuals were initially infected by HPV, but the infection subsequently cleared. We therefore classified two HPV-negative ADs, four HPV-negative SCCs, and the HPV-negative sample of unknown histology as HPV hit-and-run tumors, leaving seven ADs, five SCCs, and one rhabdomyosarcoma that were truly HPV negative. Details of the demographic and mutational characteristics of these samples are provided in **Supplemental Tables 6 and 7**.

In this cohort, we identified two cases of rare cervical sarcomas: a rhabdomyosarcoma and a leiomyosarcoma. The individual with leiomyosarcoma was 40-45 years old at biopsy, and the tumor was HPV negative. The leiomyosarcoma tumor had mutations in *B2M*, *CREBBP*, *EGR3*, *PTEN*, *SPTAN1*, and *STK11* (**Supplemental Table 8)**. The *STK11* mutation (R304W) and the *PTEN* mutation (G209fs) are known to be pathogenic. The individual with rhabdomyosarcoma was 20-25 years old at biopsy, and like the leiomyosarcoma case, the tumor was HPV negative. The rhabdomyosarcoma tumor had mutations in *DICER1*, *MGA*, *POLE*, and *RAF1*. There were two pathogenic mutations in the *DICER1* gene, D1709N and Y1546*; both were absent in the subjects’ blood DNA and, therefore, somatic mutations.

## Discussion

In the US, the five-year survival for invasive cervical cancer has improved modestly, and invasive cervical adenocarcinoma continues to have a very poor outcome [7]. Identifying key oncogenic pathways and alterations is a critical step toward subdividing cervical cancers and informing targeted therapies. This investigation has identified *STK11* as one of the most frequently altered genes in cervical cancer, specifically cervical adenocarcinoma. Alterations to *STK11* include mutations, focal and homozygous gene deletions, and whole-genome approaches are required to identify all alterations [29]. *STK11*-altered adenocarcinomas differ clinically from *STK11*-WT adenocarcinomas in several ways. Individuals with *STK11*-altered adenocarcinoma are diagnosed at a younger age than individuals with *STK11*-WT adenocarcinoma, and *STK11-*alteration is associated with poorer survival [23]. *STK11*-altered tumors are more likely to be positive for HPV18 or HPV45 and are less likely to be positive for HPV16 than *STK11*-WT tumors. However, no *STK11* alterations were detected in HPV-negative tumors. *YAP1* amplification is more common in individuals with *STK11* alterations, whereas *PIK3CA* mutations are less common and almost mutually exclusive. The combination of these clinical characteristics and the high frequency of *STK11* alterations makes *STK11*-altered tumors a distinct subtype of cervical adenocarcinomas (**Figure 6D**).

STK11, a kinase, is a known tumor suppressor. Germline mutations in *STK11* cause the autosomal dominant disorder Peutz-Jeghers Syndrome (PJS), which has an increased likelihood of several cancers, including adenocarcinoma [30-32] [33]. One normal function of STK11 is to phosphorylate and activate the AMP-activated protein kinase (AMPK) when the cell is deprived of energy [34]. AMPK then increases ATP production and conservation, promoting cellular survival but decreasing cell proliferation. Activation of the STK11/AMPK pathway also inhibits the mTOR pathway, further reducing cell growth [34]. The most common gynecological cancer among women with PJS is cervical adenocarcinoma, and more specifically, a rare form of HPV-cervical adenocarcinoma known as minimal deviation adenocarcinoma. *STK11* is the third most frequently mutated gene in sporadic lung adenocarcinoma (LUAD) and is associated with poor response to immune checkpoint inhibitors [35, 36]. In both murine models and human LUAD, *STK11* mutations are associated with lower PD-L1 expression and T cell infiltration compared to normal tissue [37].

In cervical cancer, we have shown that *STK11* mutation is associated with an earlier age of diagnosis and poorer survival, suggesting that this is a more aggressive subtype. However, little is currently known regarding the function of *STK11* in the onset and progression of cervical cancer. In the lung, *STK11* mutation or loss has been shown to result in decreased YAP1 phosphorylation, increased nuclear YAP1 abundance, and increased YAP1 signaling [38, 39]. Our data that *YAP1* amplification is more frequent in *STK11-altered tumors suggests an* interaction between these two pathways in cervical cancer. We find that *STK11* alterations are more common in HPV18 and HPV45-positive tumors compared to HPV16-containing tumors, as has been seen for other driver mutations [40].

Through whole-genome sequencing and SNP array analysis, we identified numerous focal deletions in the *STK11* gene that were not detected by WES; a similar finding has been reported in lung cancer [29]. Through analysis of gene copy number on 19p, we identified loss of the *STK11* gene region in multiple tumors, with a signature of BFB cycles [41] centromeric to *STK11*. Analysis of the whole-genome sequence identified inverted reads with staggered ends and DNA amplification, characteristic of BFB events [12]. In contrast, there are multiple examples of BFB events leading to oncogene amplification, such as *ERBB2* [42], ours is, to our knowledge, the first report of BFB associated with frequent deletion of one allele of a tumor suppressor gene.

Currently, the only FDA-approved immune checkpoint inhibitor for cervical cancer, Pembrolizumab, is more effective in cervical SCC than cervical AD [43]. Furthermore, the expression of *CD274*/PD-L1, the target of Pembrolizumab, is significantly higher in SCCs than ADs [44] **(Supplemental Figure 13**). Given the low PD-L1 expression and the limited efficacy of immune checkpoint inhibitors in STK11-mutated LUAD, Pembrolizumab may be less effective in cervical AD, in part due to the high frequency of *STK11* alterations. In the Caris sample, *STK11-altered adenocarcinomas responded poorly to ICI therapy, whereas wild-type samples showed* improved survival. Whether the effect of STK11 loss-of-function on the immune response to tumors in lung and cervical cancers is similar remains to be determined.

HPV infection causes over 98% of cervical cancer. However, it is increasingly recognized that there is a subset of cervical cancer that is HPV- or HPV-independent cervical cancers without detectable HPV infection or integration. HPV-independent cervical cancer is predominantly AD and is frequently *TP53-*mutated [28]. Our study identified five HPV- SCCs (out of 159) and seven HPV- ADs (out of 48). Half of the HPV- ADs had *TP53* mutations, but individuals with HPV- tumors did not differ in age from individuals with HPV+ tumors.

Two individuals with rare cervical leiomyosarcoma and rhabdomyosarcoma were identified in our study, both HPV-negative. The leiomyosarcoma was positive for a pathogenic *PTEN* mutation, and loss of *PTEN* expression through deletion or mutation is a known driver of soft tissue sarcomas, including leiomyosarcoma [45, 46]. The individual with rhabdomyosarcoma was the youngest individual included in our study, with an age of 20-25 at biopsy. Germline pathogenic *DICER1* variants are associated with pediatric rhabdomyosarcomas Kim, 2021 #1901}; however, both detected *DICER1* mutations in this individual appeared to be somatic, although we cannot rule out germline mosaicism.

This study represents one of the most extensive investigations of cervical cancer in LMICs and the largest study to our knowledge in Latin America, a region with a high burden of cervical cancer [1]. We attempted to validate our findings using nearly 7,000 additional cervical tumors from publicly available datasets (TCGA and AACR Project Genie) and the Caris codeAI database. The TCGA dataset is an outlier, as no individuals with adenocarcinoma had *STK11* mutations or deletions, whereas *STK11* alterations have been associated with AD in all other studies. TCGA tumors showed a similar distribution of grade to those of the Latin American cohort (**Supplemental Figure 14**). The emergence of *STK11*-alteration as a major driver of cervical cancer in Latin American subjects demonstrates the importance of using multiple patient cohorts from distinct backgrounds in identifying novel driver genes.

Our study has several limitations. Although we sequenced 306 tumors, histology was known for only 209, HPV type was known for only 229, and a matched normal sample was unavailable for some tumors. Given the low frequency of AD, we may have lacked the power to detect additional driver genes that differ between SCC and AD. The study included subjects with missing demographic data, and clinical outcomes are not available from Guatemala or Venezuela. Therefore, we were unable to investigate the association between driver gene variants and survival in these populations. Clinical outcomes data in Caris codeAI are derived from insurance claims and are incomplete for certain variables, such as tumor stage, and age is at sample collection. The Caris dataset included approximately 40% grade 4 tumors (**Supplemental Figure 14**). It will be essential to determine the functional consequences of *STK11* loss in cervical cells, particularly in adenocarcinomas.

Few FDA-approved targeted therapies for cervical cancer currently exist. Identifying novel genetic subtypes of cervical cancer is an essential step on the path to developing targeted therapies. We have previously demonstrated that *PIK3CA-*mutated and *YAP1*-amplified tumors represent distinct subtypes of cervical cancer that may uniquely respond to different treatments. In this study, we have identified *STK11*-altered tumors as an additional subtype. Given the significance of *STK11* in lung adenocarcinoma, developing specific therapies for patients with *STK11*-altered cervical tumors is imperative.

## Acknowledgements

We thank Patricia Zaid, Lineth Boror, and Esther Avila for assistance in sample collection and transportation, and Katherine Stang for comments on the manuscript. TCGA data was accessed through dbGAP. The authors acknowledge the American Association for Cancer Research and its financial and material support for the development of the AACR Project GENIE registry, as well as the members of the consortium for their commitment to data sharing. Interpretations are the responsibility of the study authors.

This research was supported in part by the Intramural Research Program of the National Institutes of Health (NIH) and the Frederick National Laboratory for Cancer Research, under Contract No. 75N91019D00024 and the Center for Cancer Research. The contributions of the NIH author(s) were made as part of their official duties as NIH federal employees, are in compliance with agency policy requirements, and are considered Works of the United States Government. However, the findings and conclusions presented in this paper are those of the author(s) and do not necessarily reflect the views of the NIH or the U.S. Department of Health and Human Services. The content of this publication does not necessarily reflect the views or policies of the Department of Health and Human Services, nor does mention of trade names, commercial products, or organizations imply endorsement by the U.S. Government.

## Notes

### Competing Interest Statement

The authors have declared no competing interest.

### Author Declarations

Ethics committees at the Instituto Nacional de Cancerologia (INCAN) in Guatemala and Hospital Central Universitario (Dr. Antonio Maria Pineda) in Venezuela gave ethical approval for this work.

